# Human Monkeypox Virus outbreak among Men who have Sex with Men in Amsterdam and Rotterdam, the Netherlands: no evidence for undetected transmission prior to May 2022 in a retrospective study

**DOI:** 10.1101/2022.11.19.22282179

**Authors:** Henry J. de Vries, Hannelore M. Götz, Sylvia Bruisten, Annemiek A. van der Eijk, Maria Prins, Bas B. Oude Munnink, Matthijs R.A. Welkers, Marcel Jonges, Richard Molenkamp, Brenda M. Westerhuis, Leonard Schuele, Arjen Stam, Marjan Boter, Elske Hoornenborg, Daphne Mulders, Mariken van den Lubben, Marion Koopmans

## Abstract

Since May 2022 over 20.000 human monkeypox virus (hMPXV) cases have been reported from 29 EU/EEA countries, predominantly among men who have sex with men. With over 1200 cases, and a crude notification rate of 70.7 per million population, the Netherlands was in the top 5 European countries most affected. The first national case was reported from May 10^th^, yet potential prior transmission remains unknown. Insight into prolonged undetected transmission can help to understand the current outbreak dynamics and aid future public health interventions. We therefor performed a retrospective study to elucidate whether undetected transmission of hMPXV occurred prior to the first reported cases in Amsterdam and Rotterdam. In 401 anorectal- and ulcer samples from visitors of the Centers for Sexual Health of Amsterdam and Rotterdam dating back to February 14^th^, we identified 2 new cases, the earliest from the first week of May (week 18), 2022. This coincides with the first cases reported in the United Kingdom, Spain and Portugal. We found no evidence of widespread hMPXV transmission in Dutch sexual networks of MSM prior to May 2022. Likely, the hMPXV outbreak expanded across Europe within a short period in the spring of 2022 in an international highly intertwined network of sexually active MSM.

**Ethical statement:** The Amsterdam University Medical Centre Ethical committee approved the study and deemed a full review not necessary according to the Medical Research Involving Human Subjects Act (reference letter: W22_257 # 22.313, dd July 14, 2022). Samples included from Rotterdam were required to be collected more than 3 weeks prior to the analysis (i.e. the duration of quarantine in case of a positive result) as they were deemed to be of no consequence for the index or his/her contacts. All samples and collected data were fully de-identified before sample analysis assuring anonymity.

## Background

Monkeypox virus (MPXV) is an orthopox virus that is closely related to the small pox virus.(1) It is endemic in Central and West African countries where it caused recurring outbreaks among humans that could be linked to zoonotic transmission. The reservoir of MPXV in the African continent are mainly rodents. On the African continent, airborne and close skin-skin contact are considered the modes of transmission, and cases develop malaise, airway complaints and a generalized monomorphic rash consisting of vesicles, pustules, and ulcers. (2) The reported mortality is between 1 and 10% depending on the virus clade and especially children and pregnant persons are affected. In the past, introductions of human Monkeypox virus (hMPXV) cases have occurred outside known enzootic (African) countries, but these events have not led to subsequent sustained transmissions. (3)

Since May 2022 after initial detection in the United Kingdom (UK), a global outbreak has evolved primarily among men who have sex with men (MSM). The transmission has mainly occurred among men with frequent sexual-, and direct skin contact with multiple partners. Genome sequencing showed that viruses from clade IIb caused the vast majority of cases. (4) The first hMPXV cases in the Netherlands were identified in retrospect on May 20^th^ 2022 in two Centers for Sexual health (CSH) based on hMPXV suspected symptoms, previously collected May 10^th^ 2022 in Amsterdam and May 19^th^ in Rotterdam. By October 2022, 20.675 confirmed cases of MPX have been reported from 29 EU/EEA countries. In the Netherlands, more than 1200 cases were diagnosed, of which >50% in Amsterdam and about 7% in Rotterdam. (5)

The exact moment of introduction and subsequent spread of sexually transmitted hMPXV among MSM is unknown. Insight into prolonged undetected transmission can help to understand the current outbreak dynamics and aid future public health interventions. Here, we tested a convenience collection of stored anorectal and ulcer samples from MSM visiting the CSH from Amsterdam and Rotterdam on hMPXV. As cut-off point, we used in retrospect the day before the first presentation of an identified case in each city, respectively May 9^th^ and May 18^th^.

## Methods

### Retrospective analysis in Amsterdam

At the CSH in Amsterdam, anorectal samples are routinely collected from MSM for *Neisseria gonorrhoeae* (Ng) and *Chlamydia trachomatis* (Ct) testing. All positive samples are stored for future reference. From visitors with anal, genital or oral (muco)cutaneous ulcers, dry samples are collected for herpes simplex virus 1 and 2 (HSV), varicella zoster virus (VZV), and *Treponema pallidum* (Tp) testing. All ulcer swab eluates are stored for 4 months. We used qPCR (4,5) to test the samples from February 14 to May 9, for the presence of hMPXV. Patient data from the electronic patient file were de-identified prior to the availability of the hMPXV test results became available.

### Retrospective analysis in Rotterdam

At the CSH in Rotterdam, routinely collected anorectal samples from MSM positive for Ng or Ct, and ulcer samples tested for HSV1, HSV2, and VZV in the period April 1 to May 18 were tested for the presence of hMPXV. For this purpose, a qPCR based either on a pan-Orthopox PCR with subsequent hMPXV detection through sequence analysis, or a monkeypox-specific target was used. (6-8) Patient data were obtained from the electronic patient file.

### Phylogenetic analysis

Whole genome sequencing was performed as described earlier (6) for hMPXV positive samples. For phylogenetic analysis all available Genbank sequences were downloaded from mPOXSPECTRUM (https://mpox.genspectrum.org). Subsampling was performed in the Nextstrain monkeypox pipeline (https://github.com/nextstrain/monkeypox) with the augur filter settings “max_date=‘2022-06-07’, sequences_per_group=‘1000’ and “–exclude-where outbreak!=hMPXV-1”. After filtering, we added the Dutch strains generated as part of this study.

## Results

In Amsterdam we tested 169 anorectal samples that were tested positive for Ct/Ng between February 14 and May 9 on the presence of hMPXV (table 1). At least 40 samples were from visitors with anorectal complaints. We also tested all 125 ulcer samples (irrespective of test result for HSV, VZV or Tp) collected from February 14 to May 9. All 169 anorectal samples tested negative for hMPXV. In the ulcer selection, we identified one hMPXV positive sample collected in the first week of May (week 18), 2022. The patient characteristics are mentioned in table 2.

**Table 1:**
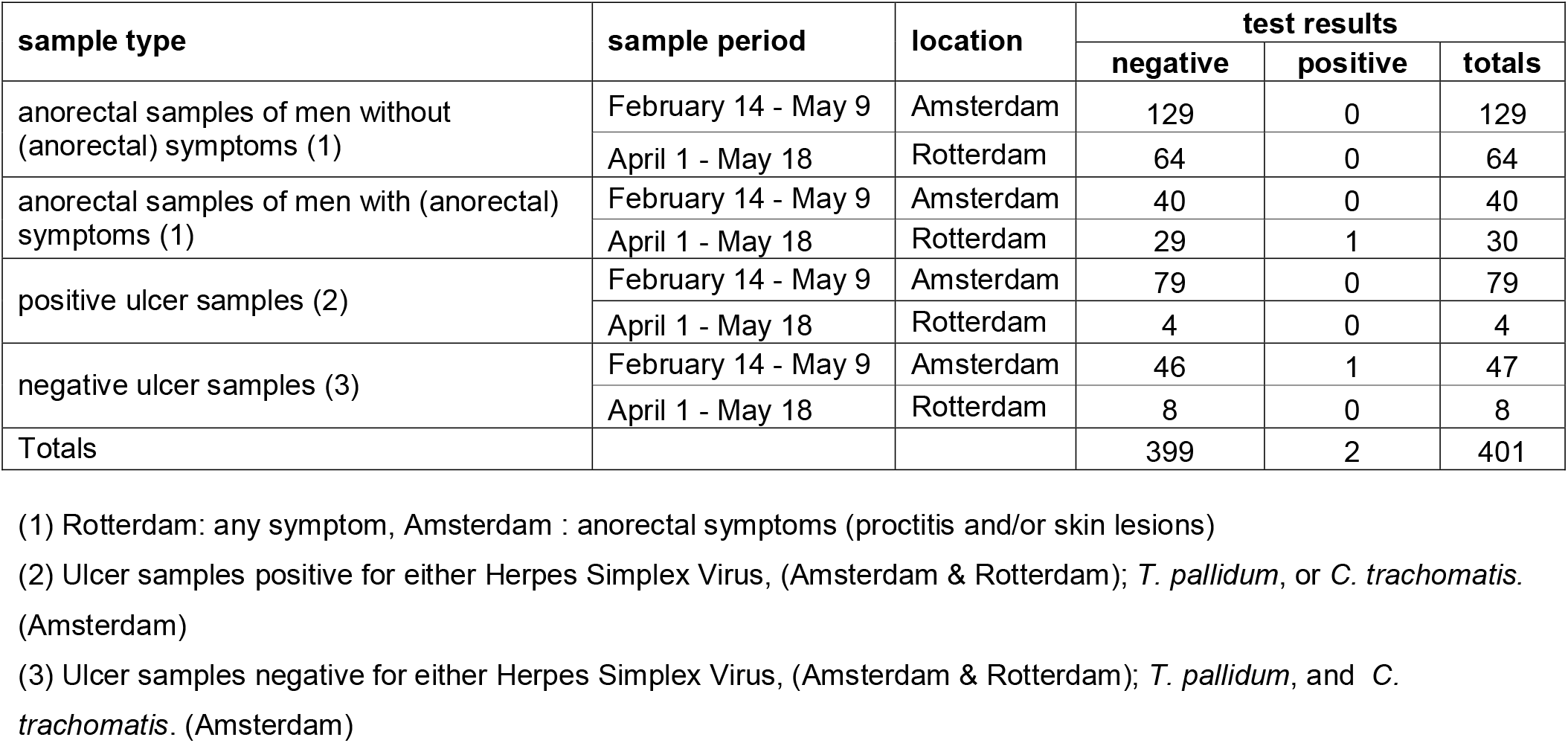
Test results for human Monkey Pox Virus of 401 samples from men who had sex with men visiting the Amsterdam and Rotterdam Centres for Sexual Health, February – May 2022.

| sample type | sample period | location | test results |  |  |
| --- | --- | --- | --- | --- | --- |
|  |  |  | negative | positive | totals |
| anorectal samples of men without (anorectal) symptoms (1) | February 14 - May 9 | Amsterdam | 129 | 0 | 129 |
|  | April 1 - May 18 | Rotterdam | 64 | 0 | 64 |
| anorectal samples of men with (anorectal) symptoms (1) | February 14 - May 9 | Amsterdam | 40 | 0 | 40 |
|  | April 1 - May 18 | Rotterdam | 29 | 1 | 30 |
| positive ulcer samples (2) | February 14 - May 9 | Amsterdam | 79 | 0 | 79 |
|  | April 1 - May 18 | Rotterdam | 4 | 0 | 4 |
| negative ulcer samples (3) | February 14 - May 9 | Amsterdam | 46 | 1 | 47 |
|  | April 1 - May 18 | Rotterdam | 8 | 0 | 8 |
| Totals |  |  | 399 | 2 | 401 |

(1) Rotterdam: any symptom, Amsterdam : anorectal symptoms (proctitis and/or skin lesions)
(2) Ulcer samples positive for either Herpes Simplex Virus, (Amsterdam & Rotterdam); *T. pallidum*, or *C. trachomatis*. (Amsterdam)
(3) Ulcer samples negative for either Herpes Simplex Virus, (Amsterdam & Rotterdam); *T. pallidum*, and *C. trachomatis*. (Amsterdam)

**Table 2:** Disease and sexual behavior characteristics of two men who had sex with men visiting the Amsterdam and Rotterdam Centres for Sexual Health who tested positive for human Monkeypox Virus infection in May 2022.

| date | location | age | symptoms | notified for STI | STI diagnosed at consultation | PrEP use | # partners < 6 months | anal condom use | groups sex | recreational drug use | Previous documented STI diagnoses |
| --- | --- | --- | --- | --- | --- | --- | --- | --- | --- | --- | --- |
| Week 18, 2022 | Amsterdam | 50-54 | multiple ulcers & itchy rash on upper legs | no | genital herpes | NA | 1 | sometimes | unknown | unknown | HIV, Ct, Ng & primary syphilis |
| Week 18, 2022 | Rotterdam | 20-24 | proctitis | yes | anorectal & pharyngeal Ct & Ng | yes | 30 | sometimes | yes | yes | yes |

PrEP: Pre exposure Prophylaxis against HIV, Ct: *Chlamydia trachomatis*, Ng: *Neisseria gonorrhoeae*, STI: Sexually Transmitted Infections

In Rotterdam, we tested 93 anorectal samples positive for Ct/Ng, of which 30 visitors reported complaints in the period April 1 to May 18 (table 1). In the same period, an additional 12 ulcer samples were also tested on hMPXV. We found one hMPXV positive anorectal sample in an MSM who reported complaints of proctitis collected in the first week of May (week 18), 2022 (table 2). All 12 ulcer samples tested negative for hMPXV.

The 2 hMPXV positive samples were obtained from the storage for phylogenetic analysis. Unfortunately, one of the hMPXV samples (collected in Rotterdam) contained too little DNA to perform successful sequence analysis. All Dutch sequences (including the additionally found strain found in Amsterdam in the first week of May, week 18, 2022) belonged to the clade IIb cluster (B.1) with a close relation to each other as well as to international strains of hMPXV (figure 1).

**Figure 1:**
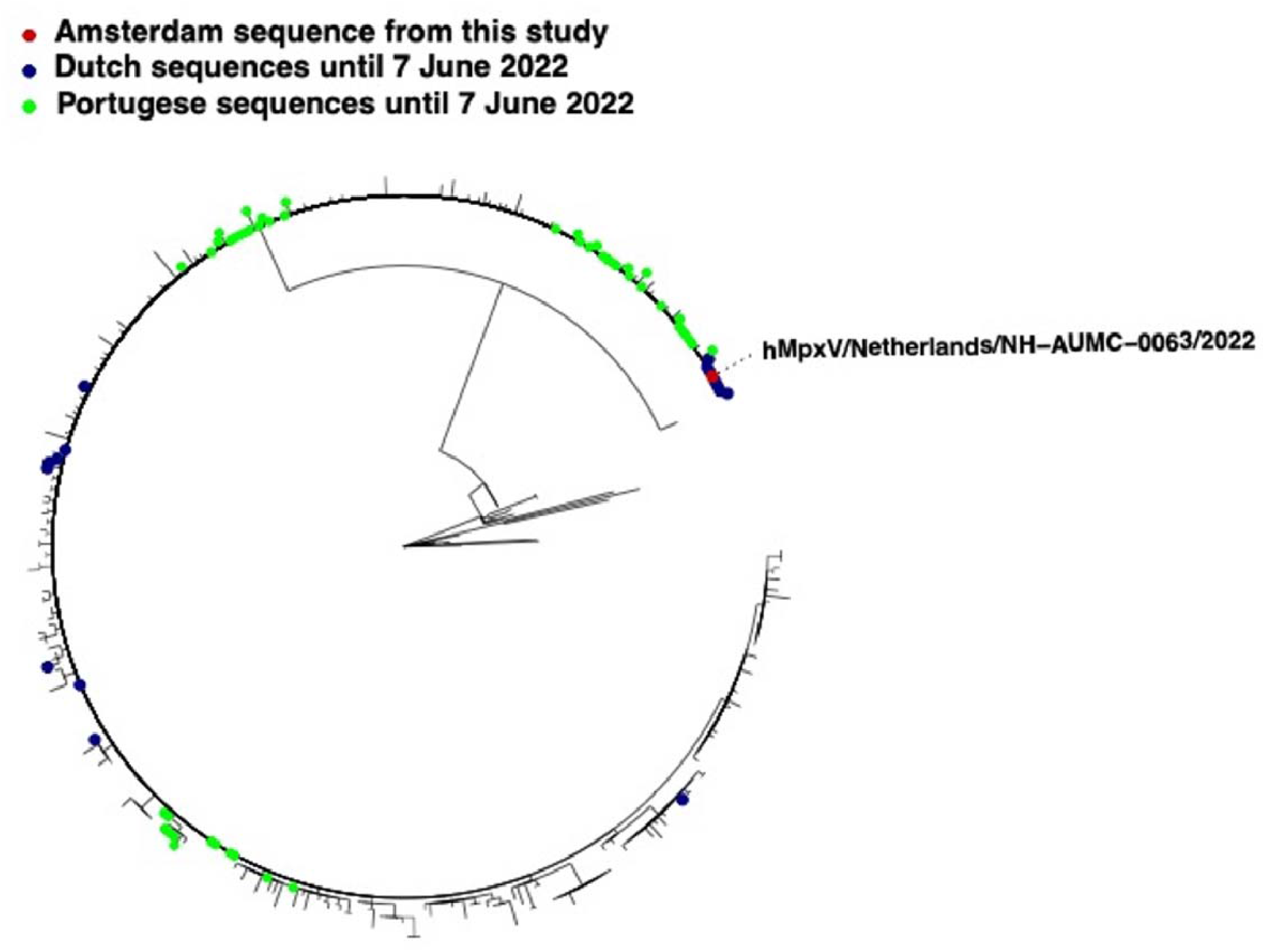
Phylogenetic association of one human Monkey Pox Virus strain found in a sample from a visitor at the Amsterdam Centre for Sexual Health in the first week of May (week 18), 2022 (red), and 11 Dutch (blue) and 48 Portugese (green) reference strains.

## Discussion

We identified two symptomatic cases of hMPXV in the first week of May (week 18), 2022 that preceded the previously first identified case in the Netherlands. We found no hMPXV in 399 samples predating the first cases. Our study included samples from 208 MSM with complaints suspected of hMPXV infection according to the current WHO suspected case definition criteria, such as anogenital ulcers (n=138) and proctitis (n=70). (9) We therefore assume that it seems unlikely that substantial undetected transmission of hMPXV occurred in the Netherlands before the first week of May (week 18), 2022. The mean incubation period for cases identified in the Netherlands was 8.5 days [4.2 to 17.3]. (10) These findings suggest that the introduction of hMPXV in Dutch sexual networks of MSM started somewhere at the end of April 2022. This coincides with the earliest symptom onset of hMPXV cases in the UK on April 21 (11, 12), in Spain on April 26 (13), and in Portugal on April 29. (14) In combination with the phylogenetic analysis of hMPXV genome sequences, showing that the initial cases across Europe are clonal, it is likely that the hMPXV outbreak expanded internationally within a short period (weeks) in the spring of 2022 in an international highly intertwined network of sexually active MSM. The outbreak also coincided with international relaxation of coronavirus disease (COVID-19) prevention measures, and resumption of global travel. (15) Both cases, had characteristics that are associated with increased risk of spread of sexually transmitted infections such as: multiple previous STI, multiple sex partners, condomless (anal) sex, either HIV PreExposure Prophylaxis use or living with HIV and substance use. These characteristics predispose for Sexually Transmitted Infections and for hMPXV. (3, 16)

The strength of our study is that we had an extensive number of lesional and anorectal samples available from persons that meet the current case definition for hMPXV. However, a limitation is that the group tested is biased as the anorectal samples were only stored if they had tested positive for Ct and/or Ng. Also, due to privacy regulations the study was anonymous. Hence, we could not gather additional data on date of symptom onset, recent sexual partners, and travel history.

In the current outbreak in MSM, it remains unclear how hMPXV was introduced. Apart from network effects (17), the currently circulating strains differ from previous hMPXV strains by over 50 nucleotides. It remains unclear whether such changes led to increased (sexual) transmission. Further viral characterization is needed as well as continued genomic surveillance on circulating hMPXV strains. (4) In conclusion, we found no indication of extensive undetected transmission of hMPXV among MSM in Amsterdam or Rotterdam before May 2022. This is in accordance with findings from other European cities and in support of a clonal international outbreak in the spring of 2022.

## Data Availability

All data produced in the present work are contained in the manuscript

https://mpox.genspectrum.org

https://github.com/nextstrain/monkeypox

## Data availability

All Dutch sequences are publicly available via Genbank and GISAID (accession numbersEPI_ISL_13728303, EPI_ISL_14752293, EPI_ISL_15641541-EPI_ISL_15641590).

## Authors contributions

In Rotterdam HMG, MK, AvdE, RM initiated the study, and HMG collected the epidemiological data. In Amsterdam HdV, SB, EH, MP and MvdL initiated the study, and SB, DM, AS, MRAW, and BW performed the laboratory tests and analyses. BOM, MB, LS, MJ and MRAW performed whole genome sequencing and phylogenetic analysis. MRAW and RM were also involved in laboratory diagnostics. All authors were actively involved in drafting, reviewing and final approval of the manuscript.

